# More than the eye can see – shedding new light on SARS-CoV-2 Lateral Flow Device-based immunoassays

**DOI:** 10.1101/2021.03.06.21253048

**Authors:** Garrit Koller, Alexander P Morrell, Rui Pedro Galão, Suzanne Pickering, Eithne MacMahon, Joanna Johnson, Konstantin Ignatyev, Stuart JD Neil, Sherif Elsharkawy, Roland Fleck, Pedro Miguel Pereira Machado, Owen Addison

## Abstract

Containing the global SARS-CoV-2 pandemic has been an unprecedented challenge due to high horizontal transmissivity and asymptomatic carriage rates. Lateral Flow Device (LFD) immunoassays were introduced in late 2020 to detect SARS-CoV-2 infection in asymptomatic or pre-symptomatic individuals rapidly. Whilst LFD technologies have been used for over 60 years, their widespread use as a public health tool during a pandemic is unprecedented. By the end of 2020, data from studies into the efficacy of the LFDs emerged and showed these point-of-care devices to have very high specificity (ability to identify true negatives) but inadequate sensitivity with high false-negative rates. The low sensitivity (<50%) shown in several studies is a critical public health concern, as asymptomatic or pre-symptomatic carriers may wrongly be assumed to be non-infectious, posing a significant risk of further spread in the community. Here we show that the direct visual readout of SARS-CoV-2 LFDs is an inadequate approach to discriminate a potentially infective viral concentration in a bio-sample. We quantified significant immobilized antigen-antibody-label conjugate complexes within the LFDs visually scored as negative using high-sensitivity synchrotron X-ray fluorescence imaging. Correlating quantitative X-ray fluorescence measurements and qRT-PCR determined numbers of viral copies, we identified that negatively scored samples could contain up to 100 PFU (equivalent here to ∼10,000 RNA copies/test). The study demonstrates where the shortcomings arise in many of the current direct-readout SARS-CoV-2 LFDs, namely being a deficiency in the readout as opposed to the potential level of detection of the test, which is orders of magnitude higher. The present findings are of importance, both to public health monitoring during the COVID-19 pandemic and to the rapid refinement of these tools for immediate and future applications.

## Introduction

Containing the global COVID-19 pandemic has been an unprecedented challenge due to high horizontal transmissivity and asymptomatic carriage rates. Tests to rapidly detect SARS-CoV-2 infection in asymptomatic and pre-symptomatic individuals in the mass population were introduced in late 2020 and are considered essential to enable people to gather safely in workplaces, educational settings and socially while maintaining control over virus transmission. However, such testing programmes are expensive and in the USA alone could cost an estimated $11-34 bn^1^ annually. The most accessible and logistically scalable diagnostic test technologies are Lateral Flow Devices (LFDs) which have been used for over 60 years as facile immunoassays^2^. However, their use during the current COVID-19 pandemic is the first mass deployment of LFDs to address a significant public health crisis. The qualitative readout of LFDs relies on the the immobilization of chromogenic complexes within a test capture field. In most LFD designs, a diluted biological sample is deposited onto a nitrocellulose substrate and wicks laterally down a test strip. A positive test is indicated when a visually discernible line develops due to the deposition of sufficient antigen-bound nanoparticle-antibody complexes within the test field containing a secondary antibody. Of the nanoparticle labels available, gold has been used extensively due to its flexibility for functionalization and is currently employed in several of the most widely used SARS-CoV-2 LFDs. However, the reliance on visual signal perception introduces variability in reported LFD performance from different sampling or analysis settings^3^. This is particularly notable when low quantities of complexes are present in a test field. Therefore, alternative approaches have been proposed to improve the sensitivity of LFD readouts, such as the use of laser-based detection systems^4^ or signal amplification using nanozyme-mediated chemiluminescence^5^. Still, these limit the usefulness to a broader population and have had limited uptake in the COVID-19 pandemic.

Much hope was placed in making LFD-based tests widely available and administered without significant training and infrastructure. However, by the end of 2020, clinical testing data from studies into the LFD efficacy emerged^3, 6-9^ and demonstrated the disparity between manufacturer-reported and results obtained clinically in mass testing. These results indicated an ‘in-field’ sensitivity of as low as 48.9% and missing as many as 23 of 45 qRT-PCR-confirmed positive cases, however, did demonstrate the tests had high specificity at >99.9%^3, 10^. Findings of inadequate sensitivity have since been reported in for many SARS-CoV-2 LFDs from different manufacturers, with few passing national benchmark tests^8, 10^. Notably, non-linear trends between the qRT-PCR *Ct*-value (cycle threshold –which provides an estimate of viral load) and false-negative rates were observed in numerous studies^3, 7, 8, 11^. High PCR *Ct*-values, suggesting low viral titres, were most likely to lead to detection failure, which has implications on viral transmission risk assessment^12^.

A public health concern is the spread of the SARS-CoV-2 virus by asymptomatic or pre-symptomatic individuals who may have an increased sense of confidence provided by a false-negative LFD result^13^. We hypothesize that the limited sensitivity of direct readout LFDs may be attributable to the failure of generating a signal (visual readout) due to the low density/mass of the immobilized reporter label, rather than the inability to bind to antigen at low titres or background signal. Here, we test this hypothesis using high sensitivity X-ray-based imaging with synchrotron light to quantify antigen-antibody-label conjugates within ‘negative’ result LFD tests.

Given the high importance placed in de-centralized testing for viral presence or infectivity, even moderate gains in signal intensity of these assays would dramatically improve control of spread and public health models guiding policy. This would stand particularly true if these enhanced readouts can be attained through readily accessible technologies with sensitivity to lower viral titres without introducing decreased specificity of the tests. As these tests exhibit high specificity, there is a potential opportunity to amplify the readout signal to increase sensitivity, given the low rates of false-positive results recorded in field trials ^3, 10^. We demonstrate in contrived, spiked tests (below the visual detection threshold) that these tests have the intrinsic capacity for lower detection limits.

## Materials and Methods

### Preparation of viral stocks and inactivation

An original stock of SARS-CoV-2 England 02/2020 strain (Public Health England, Colindale, UK) was expanded in Vero E6 cells (African green monkey kidney cells) maintained in Dulbecco’s minimal essential medium (DMEM) containing 10% foetal bovine serum (FBS) at 37°C with 5% CO2. Supernatants were harvested 72 hours post-inoculation, aliquoted and stored at -80°C. Titres were determined by plaque assays, in which 10-fold serially diluted stocks were applied to Vero E6 cells and incubated for 1 hour at 37°C. An equal volume of a pre-warmed overlay (0.1% agarose in DMEM supplemented with 2% FCS) was then added to each well, and cultures were incubated for 72 hours as above, before fixation with 4% formaldehyde and stained with 0.05 %(w/v) crystal violet in ethanol. As a surrogate for infectivity within a given target cell line, viruses propagated *in vitro* are quantified by measuring the plaque-forming capacity of serial dilutions of supernatant, measured in plaque-forming units (PFU). To generate non-infectious heat-inactivated (HI) particles, viral stocks were serially diluted, plaque-forming units were enumerated (PFU) and heat-treated at 70 °C for 30 minutes. Inactivation was confirmed by Vero cell culture, subjected to serially diluted and heat-treated viral particles.

### Quantification of SARS-nCOV2 by qRT-PCR

RNAs were extracted from 100ul of serially diluted SARS-CoV-2 stocks with pre-determined PFU inputs, using the Qiagen QIAamp Viral RNA Kit, following the manufacturer’s instructions and final eluted in 60µl of water. RT-PCR reactions were performed with 5µl of eluted RNA, 4x TaqMan Fast Virus 1-Step Master mix (Applied Biosystems) and CDC’s IDT Primer-Probes Sets targeting SARS-CoV-2 N gene regions or human RNAse P, using a QuantStudio 5 qPCR machine (ThermoFisher Scientific). RNA standards were extracted as above from serial dilutions of a NATtrol™ SARS-CoV-2 Stock (ZeptoMetri) of known RNA viral load.

### Spiking of LFD immunoassays

Two Public Health England-approved SARS-CoV-2 Antigen LFDs (Innova Medical Group Inc, Monrovia, CA, USA) and SureScreen (SureScreen Technologies Ltd, Derby, UK) were used in the present study. The former has been widely used in the clinical and asymptomatic screening of health workers, with superior performance to other LFDs submitted for approval. The contrived test samples were prepared by spiking LFD extraction buffer with serially diluted heat-inactivated or live SARS-CoV-2 virus ranging from 0 - 10,000 PFU/test. The second SARS-CoV-2 LFD system (SureScreen) was spiked with equivalent doses of heat-inactivated virus only and used as a control/comparator for visual scoring. A qualitative optical readout was performed as per the manufacturer’s instructions under identical lighting conditions for each test by multiple assessors and binary and visual-analogue score for intensity recorded.

### Optical microscopy of LFDs

LFDs were optically imaged using a Keyence VHX-7000 digital microscope (Keyence Corp, Osaka, Japan). Imaging variables were consistent between tests, including lighting (full ring), magnification (x20) and image exposure times (20 ms). LFDs were imaged at similar times after deposition of the sample. Images were aligned and cropped for visualization. Image data from all heat-inactivated Innova LFDs were subsequently stacked and converted into a single 8-bit grayscale image. Image data were combined to prevent any discrepancies between samples attributed to RGB to greyscale conversion or data compression. Test regions on each LFD were isolated, and line profiles were extracted by averaging the image along the y-axis. The resulting profiles were subsequently background fitted and corrected to alleviate variations between LFD intensities caused by different white balances, likely due to trace moisture remaining.

### Preparation of LFDs for µXRF imaging

After allowing to air dry, the test kits were disassembled and the nitrocellulose test strips mounted onto ultrapure fused silica slides (Spectrosil 2000, Heraeus Quarzglas GmbH, Kleinostheim, Germany), covered with 40 µm thickness Kapton (DuPont Inc, Wilmington, DE, USA) before high sensitivity X-ray fluorescence (XRF) imaging. Only heat-inactivated Innova samples were prepared for X-ray measurements.

XRF measurements were conducted at the microfocus spectroscopy beamline I18 at the Diamond Light Source (Harwell, UK)^14^. After initial scouting acquisition maps, 1800 x 180 µm (h x v) regions were imaged through the test and control zones of the nitrocellulose LFD strips, using a 5 x 5 µm beam footprint, a 5 µm horizontal and vertical step-size and a 400 ms acquisition time in air. An incident X-ray energy of 12.5 keV was used to result in Au L3M5 fluorescence emission yield probabilities of ∼70%. Two 4-element Vortex Si drift detectors (HitachiHi-Technologies Science, Tokyo, Japan) were positioned at 45° to the sample, and measurements were carried out at room temperature. The fluorescence signal from a homogeneous metal film reference standard (AXO, Dresden, Germany) was measured at the same geometry as the test samples to quantify the gold signal. Quantitative calculations were performed in software PyMCA using inbuilt fundamental parameter algorithms ^15^.

The extracted Au images from each LFD dilution were subsequently aligned and cropped. The image data displayed is on a common scale (95^th^ percentile), allowing for a direct, visual comparison. A logarithmic scale is used to aid visualization as a high variability of Au concentration was observed between tests. Line profiles were extracted from each Au image by averaging pixels along the y-axis. Average Au values are reported, which reflect the average mass fraction within the test pad region of each LFD. Maximum values were also extracted from the test region and represent the maximum Au concentration within a single pixel (5 x 5µm).

### Transmission Electron Microscopy

Positive control regions from nitrocellulose LFD strips were sectioned and mounted in embedding resin, Embed 812 (EMS, Hatfield, PA, USA), polymerized for 24 h at 60 °C. Ultrathin sections (70 nm) were cut using a UC7 ultramicrotome (Leica Microsystems GmbH, Wetzlar, Germany) and collected on formvar /carbon-coated slot grids. In parallel, gold nanoparticles were eluted into ddH2O with sonication and centrifuged to separate. For imaging, particles were diluted 1:1 in ddH2O and adsorbed on carbon film copper grids for 5 min. Samples were imaged using a TEM operated at 120 kV (JEOL JEM 1400Plus, JEOL, Tokyo, Japan) equipped with a 2000 x 2000 format CCD camera (JEOL Ruby CCD Camera, JEOL, Tokyo, Japan).

## Results and Discussion

The rapid development and implementation of assays for detecting SARS-CoV-2, by necessity, involved exploiting existing technologies with a limited time for thorough pre-market validation. Whilst applicable to all diagnostic platforms for SARS-CoV-2, including RT-qPCR based assays where initial protocols reported PCR-negative findings in many as 40% of biological specimens from known infections16, this was particularly applicable to LFDs^3, 6, 8, 11, 17^. Concerns were further raised following reporting of post-deployment surveillance studies of LFDs that concerns about test sensitivity have arisen, and many were quickly described as ‘not fit for purpose’ for identifying SARS-CoV-2 positive patients ^6, 18^. However, some observers suggested a more nuanced view on LFD utility, particularly as more insights are emerging on what constitutes infectious doses^10^, further supporting the potential public health value of LFD technology in mass testing.

Identifying approaches that can discriminate between the various contributing factors in assay design that combine to dictate the sensitivity and specificity of SARS-CoV-2 tests is an unsolved problem that needs urgent attention^12^. Insights gained from addressing these issues will likely aid future LFD development, including its additional role as a public health tool. To the authors’ knowledge, the present study is the first report of using synchrotron XRF-based detection to delineate the maximum achievable detection thresholds of these devices. By correlating different approaches, the signal limit of detection for each method can then be compared with the ‘gold standards’ of infectious dose (PFU) and corresponding diagnostic RT-qPCR-based quantitation.

Here we first demonstrate good agreement between the Innova LFD results spiked with infectious and heat-inactivated virus, with a consistent threshold of visual detection observed at a viral concentration between 20 and 100 PFU **(Figure 1a)**. The findings of near-parity in infectious and heat-inactivated viral particles across platforms are critical from a technology-development perspective, facilitating the method development and validation within biosafety level (BSL) I, rather than BSLIII, facilities, without appreciable decreases in assay performance at various PFU-levels. The finding of viral RNA being conserved in a similar manner^19^ for heat-inactivated samples may enable combined nucleic acid detection in tandem with antigen enrichment.

**Figure 1.**
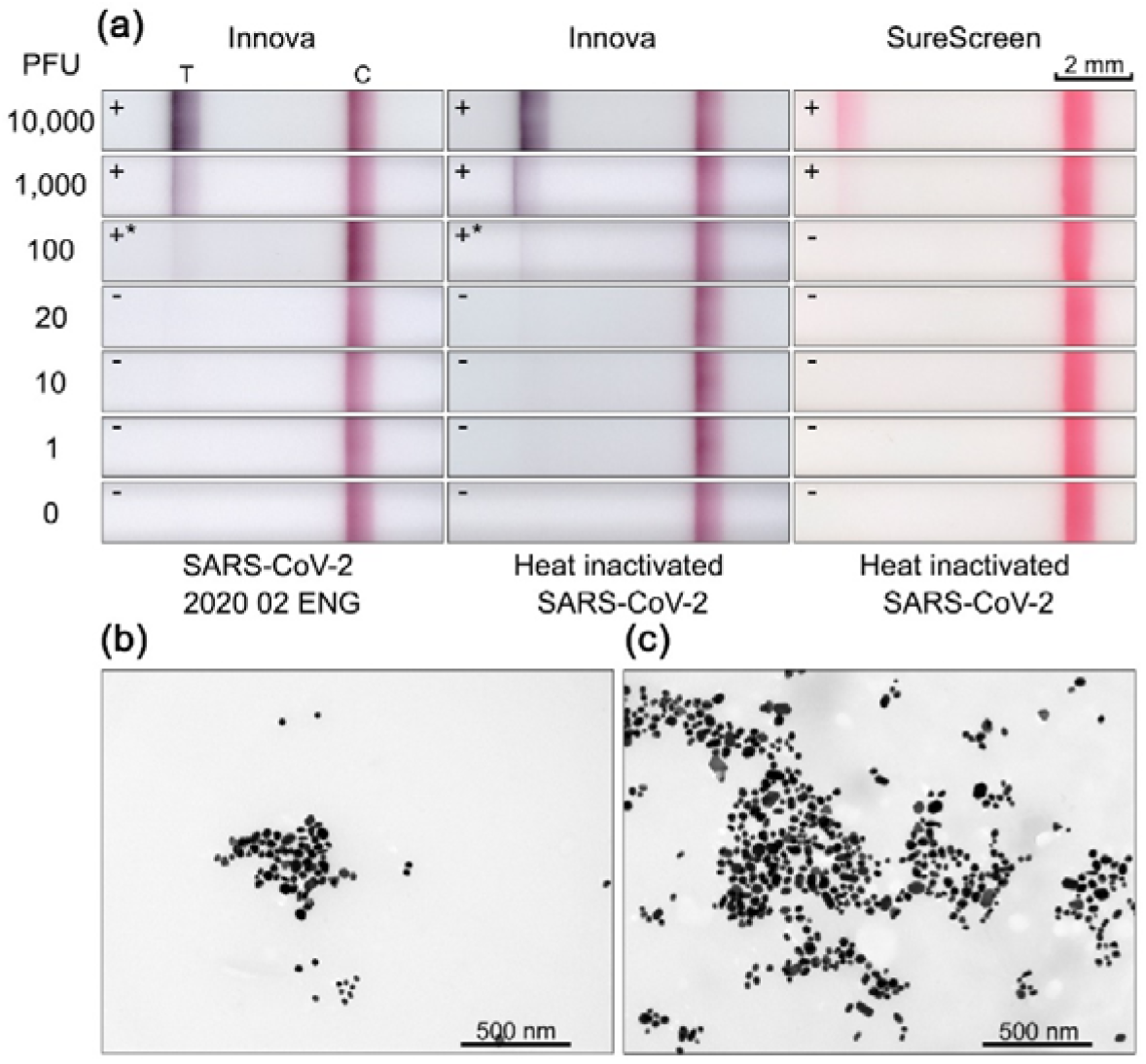
**(a)** Digital microscopy images of LFD strips from Innova (left and middle) and SureScreen (right) models. An excellent visual and performance agreement was observed between the live and the heat-inactivated virus. The SureScreen offered marginally lower visual sensitivity compared with the Innova tests. **(b)** and **(c)** TEM images of Au nanoparticles within Innova LFD immobilized on the nitrocellulose strip and subsequently eluted from the LFD, respectively. Labels‘ T’ and ‘C’ refer to the test and control regions on the LFDs, respectively.

In keeping with the previous reports^9^, the two LFDs (Innova and SureSreen) had different visual detection thresholds. In both cases, no signal was observed in spiked tests at <100 PFU. The Innova LFD uses immunolabelled gold nanoparticles as its core technology, which can be seen in **Figure 1b** on the nitrocellulose test strip either as visually detectable agglomerations of ∼500 nm or as visually undetectable dispersed single particles. The blue-black readout for the Innova LFD on the test zone (compared with the control zone) may be explained by surface plasmon resonance arising from approximating antigen-bound gold nanoparticles.

Taken together, the data suggest that visual detection requires optimization of the nanoparticle agglomeration density and the surface area coverage within the test strip zone, both of which are affected by the individual nanoparticle size. Nanoparticles eluted from the Innova LFD **(Figure 1c)** exhibited a limited size range from ∼40-60 nm. Thus, quantifying by mass provides a correlation with the numbers of antigen-antibody complexes present. The common use of immunolabelled gold nanoparticles in LFD also presents a unique opportunity for quantitative measurement using XRF over several orders of magnitude of viral titer to demonstrate avidity and binding modes and background signal arising from non-specific deposition.

Results correlating the mass fraction of gold across the test strip with viral PFU counts are summarised in **Figure 2. Figure 2a** shows the standardized imaging regions for XRF within the Innova LFD strip across conjugate areas. **Figure 2b** shows, on a common scale, the mass fraction and pattern of gold nanoparticles detected. As a surrogate indicator of antibody affinity and avidity for the complex deposited, a distinct pattern of gold distribution is observed within the test zones, with an abrupt amplitude transition to high-level Au deposition/signal and a gradual decrease to baseline in a dose-dependent manner, albeit becoming increasingly heterogeneous with decreasing viral PFU. In **Figure 2c**, the average mass fraction per spiked test viral PFU is reported and shows that all tests have elevated detectable gold within the test zone compared with the LFD run with no activation of the conjugate material (labelled E). The average mass fraction of gold for a test spiked at the lowest concentration (1 PFU) exhibited a threefold greater average mass fraction than the extraction buffer control sample (4.22 vs 1.40 ×10^−7^). The majority of the signal was distributed within the test zone. Notably, for all ‘dilute’ samples (<100 PFU), the maximum pixel intensity was similar and is likely a result of a consistent agglomeration phenomenon and a similar number of nanoparticles associating with an antigen. This observation is important concerning potential strategies for better discriminating a readout, demonstrating different densities observed relative to the unbound control sample. This is exemplified by using gold nanoparticle conjugates dually absorbed with antibodies and use of a secondary reporter, horseradish peroxidase (HRP), enabling further development after the initial LFD run has completed (Supplementary methods & Supplementary Figure 1).

**Figure 2.**
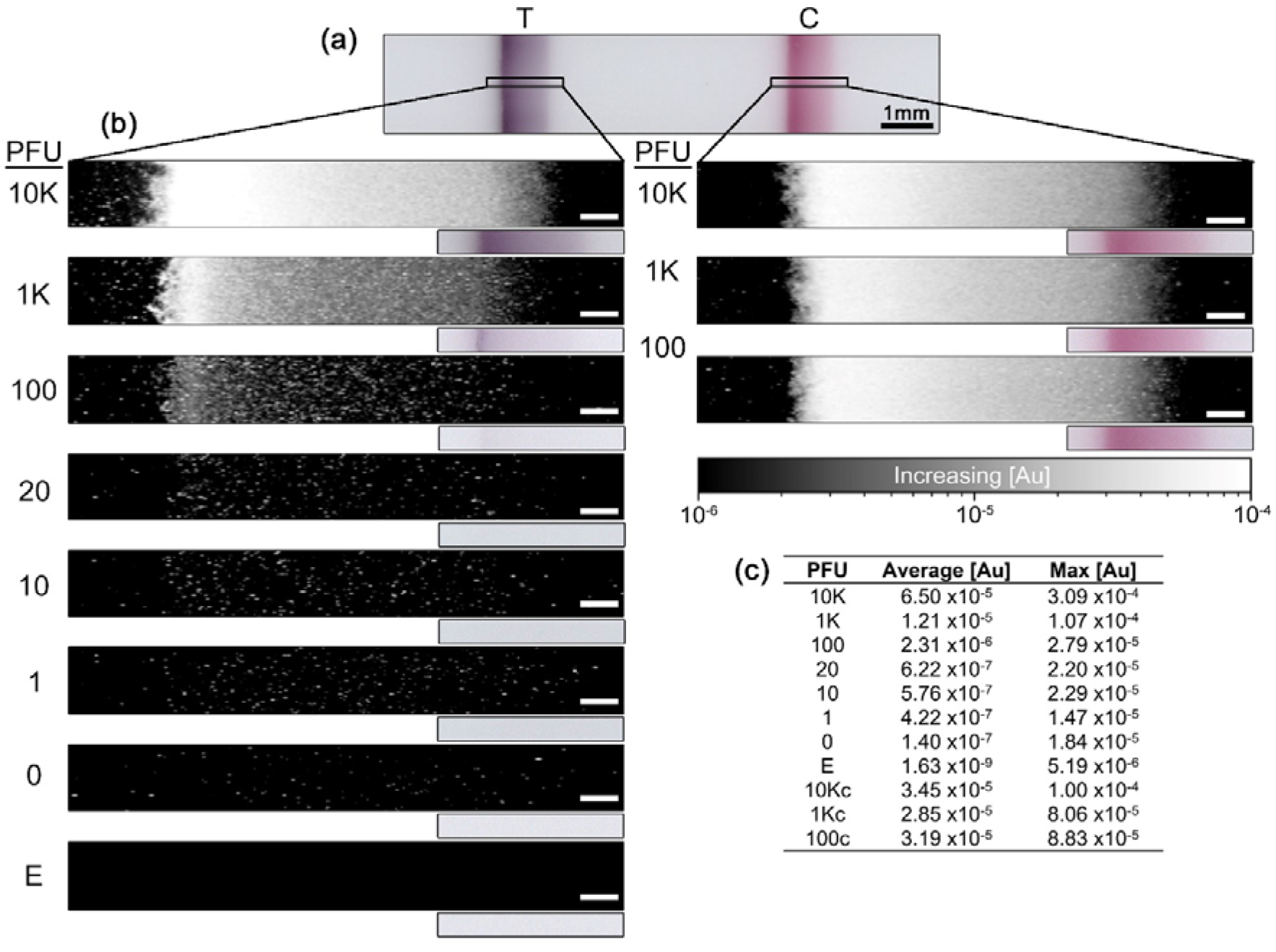
Mass fractions for viral dilution series **(a)** Digital microscopy image highlighting regions of interest within LFDs selected for subsequent synchrotron XRF measurements. **(b)** Au XRF images, expressed as mass fractions, on a common logarithmic scale are shown from the test and control strips from LFDs with descending PFU values. Below each XRF image shows the corresponding optical image within the same region of interest. **(c)** a summary of quantitative values extracted from synchrotron XRF images, including average and maximum values within the image data.

In **Figure 2a**, a scattered 2D distribution of bright spots (high gold fluorescence) can be seen, with incomplete but consistent, presence across a substrate. An analogy can be drawn with the ‘Ben-Day’ dot printing approach, which was the hallmark of the 1960s American artist Roy Lichtenstein, where varying the density of identically sized and coloured dots generates tonal range detectable by the human eye. Through simple control of the density of ‘spots’ even in low numbers, contrast can be achieved. The potential of such approaches is illustrated here in the XRF map of the 100 PFU spiked LFD, where variability in clustering density **(Figure 2b)** correlates with visual contrast difference seen in **Figure 1a**. A simulation of increasing the density of a signal is shown in **Figure 3**. The density of the detected antigen-antibody complex was increased without altering signal intensity or total number of pixels with sufficient XRF signal for a 20 PFU spike test (**Figure 3a,b**). This provided a concentrated band with sufficient average mass fraction of Au that would be visibly detectable (**Figure 3c,d**).

**Figure 3.**
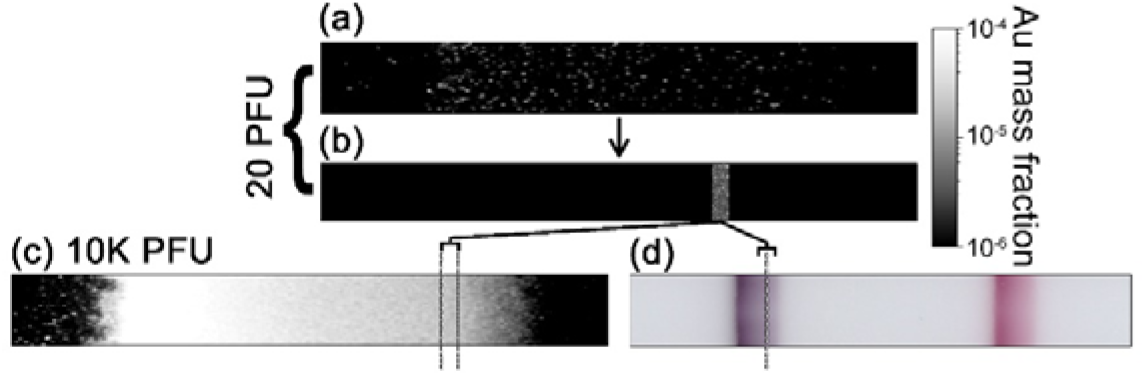
**(a)** Quantitative XRF Au image of 20 PFU spiked LFD across the test zone. (b) Simulation of a test zone where pixels from **(a)** were condensed into a contrived band. The resultant concentration and density of the simulated image would now be visibly detectable. This is demonstrated by comparing the XRF **(c)** and optical image **(d)** for the 10K PFU test.

Plots of the average gold mass fraction for each viral exposure PFU across the test zone are shown in **Figure 4a,b** and summarised in **Figure 4c**, where a log-linear trend between increasing mass fraction of gold and PFU from 1 -100 is observed, highlighting a high signal-to-noise ratio. This apparent pattern (R^2^ =0.98) gives confidence in the potential sensitivity of the test when used with dilute biological samples and further supports that efforts for developing methods for signal amplification should be made. Similar analysis approaches were subsequently applied to standardized optical imaging of the LFD tests. Surprisingly, even at extremely low pixel intensity for low viral counts (1-20 PFU), a signal pattern similar to that observed for XRF data was detected **(Figure 4d-f)**. This finding demonstrates the potential utility of simple optical means of increasing detection sensitivity and may guide further work on minimal detection signal frequency and amplitude across a test field required to provide robust readouts. No matrix effect on signal intensity and deposition pattern was observed in buffer-only or contrived samples (Supplementary Figure 2). Taken together, the observation of dose-dependent effects, demonstrated in previous reports^8^, can in large parts be accounted for by the inability to discriminate weak signals irrespective of matrix effects. However, further studies would be warranted to examine the potential contribution to readsout arising from changes in the biological matrix, host-derived antibody titers such as lymphocytoplasmic exudates, concomitant superinfections and within different symptomatic presentations, and thus represent a shortcoming of the present study.

**Figure 4.**
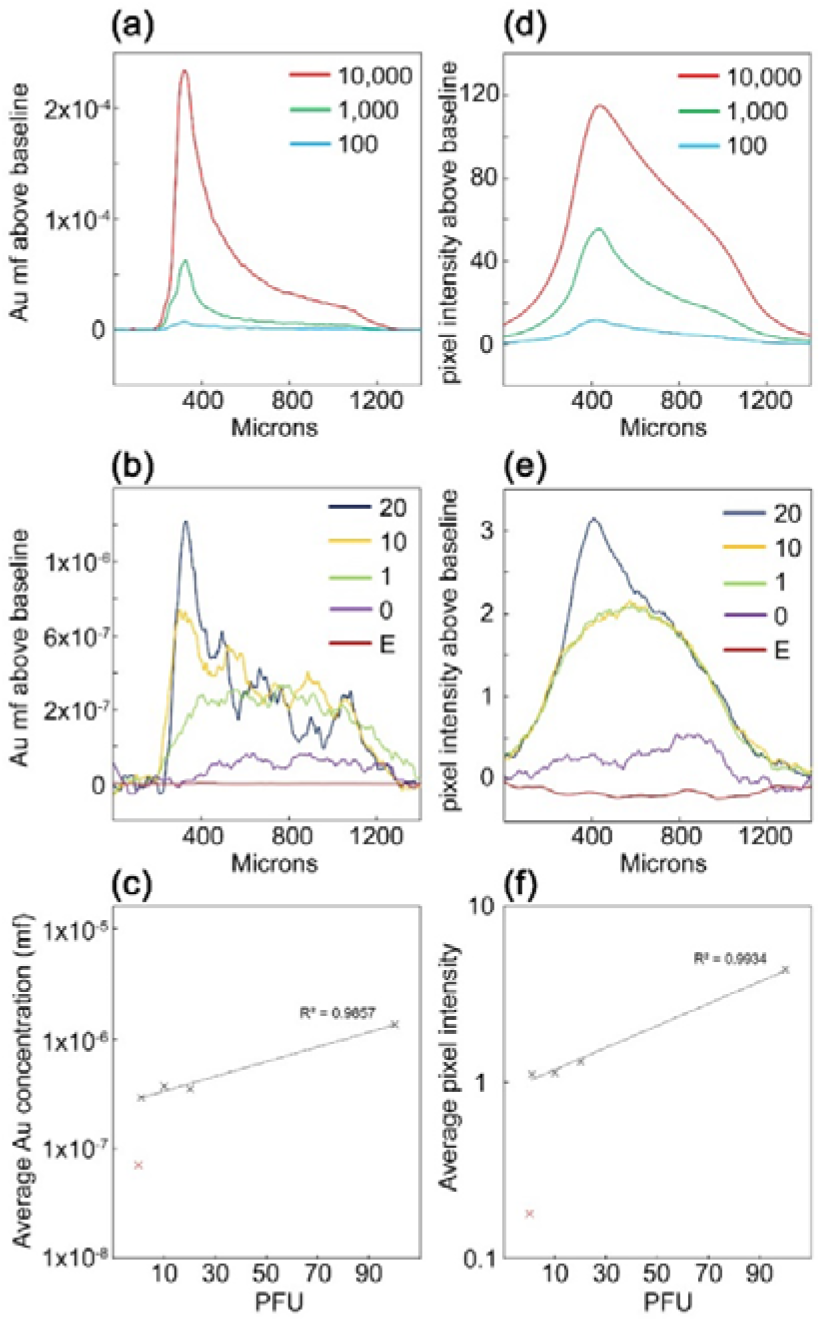
**(a)** and **(b)** Line traces extracted from XRF LFD images are shown. **(c)** Displays the log-linear trend between the average Au concentration acquired from XRF data and the respective PFU applied to the LFD. **(d)** and **(e)** line traced extracted from the optical microscopy images are shown. **(f)** Displays the log-linear trend between the average pixel intensity acquired from the optical microscopy data and the respective PFU applied to the LFD.

A threshold for subjective detection was found to lie between 20 and 100 PFU for the LFDs examined. This approximates the cycle threshold observed for previous studies correlating viral numbers from qPCR cycle numbers and the ability to form cytotoxic plaques in cell culture^20-25^. In this study, RT-qPCR validation demonstrated that 100 PFU/test was equivalent to ∼10,000 RNA copies/test and a *Ct* value of 27. Whilst there is no exact measure of how many viral particles are considered infectious, the gold standard of confirming and quantifying the ability of SARS CoV-2 to infect is a culture-based approach, with some studies demonstrating infectivity, as measured by cytopathic effects in a dose-dependent manner^20, 22, 26-29^. Several studies have conducted side-by-side comparisons between the qRT-PCR *Ct* value of a clinical specimen and viral load, establishing linear relationships between the *Ct* value of samples and the corresponding infectivity ascribed using culture-based assays. Given the high degree of correlation between detection modalities within our study, we demonstrate that the improvements with simple, optical-based methods may enable reduced false-negative rates. This is particularly true for viral loads <100 PFU/test, and improvements in this range may help define an infectious dose detection threshold. The advantage of such optimized LFDs over RT-qPCR detection is that they are not confounded by the shedding of residual viral RNA that may yield positive results for several weeks after resolution of the infection. Patients with prior SARS-CoV-2 infection may have positive PCR results in samples taken many weeks afterwards due to the presence of detectable RNA at high *Ct* values but do not harbour the infectious virus. This is a real challenge for infection control policies and has led to the policy of avoidance of testing anyone with known infection in the previous 90 days. In some way, the lesser sensitivity of SARS-CoV-2 LFD tests has been viewed as advantageous if it can be correlated with infectiousness.

Furthermore, it must be sensitive enough to identify infectious positives, which is the current deficiency of current designs. The optimal LFD for SARS-CoV-2 would therefore be less sensitive than PCR testing, however, sensitive enough to detect the virus in quantities that are likely to be infectious. This is the clear technological development need for future SARS-CoV-2 LFD assays.

## Conclusions

The present study highlights significant weaknesses of currently used LFD devices, potentially generalizable, mechanistic insights into the activation of the test zones, and providing the basis for improved SARS CoV-2 detection through increased signal amplification. Although synchrotron-based XRF approaches are not a readily accessible means of validating assays, the data produced demonstrates that the LFD technology platform can predictably immobilize viral antigen from dilute specimen enables improvements of readout signal without increasing false positivity. The results delineate theoretical detection thresholds and the contribution of signal-to-noise or unspecific events affecting signal interpretation, with hundred-fold improvements potentially possible. Attaining these improvements would match and likely surpass the resource-intensive RT-qPCR methodology. Furthermore, it highlights that, despite the plethora of proposed methods published to improve point of care test sensitivity and specificity, few of these have found widespread application within this decade-old but tried and tested diagnostic modality.

## Data Availability

Associated data is contained within the document.

## Acknowledgements

The authors gratefully acknowledge the rapid access beamtime session SP28216 awarded at the Diamond Light Source. The following reagent was obtained through BEI Resources, NIAID, NIH: Monoclonal Anti-SARS-Related Coronavirus 2 Spike Glycoprotein (IgG Mab, Rabbit), NR-53788.

