## Supplementary materials for "More than the eye can see – shedding new light on SARS-CoV-2 Lateral Flow Device-based immunoassays"

**Supplemental Materials**

**Methods**

*Horseradish peroxidase-functionalised gold nanoparticles targeting anti-SARS-CoV-2 spike protein*

A suspension of 10 nm colloidal gold nanoparticles (O.D._520 nm_ = 1; BBI Solutions, Crumlin, UK) was adjusted to pH 9 with NaCO_3_ and incubated with 3 µg / mL rabbit monoclonal antibody targeting SARS-CoV-2 Spike Glycoprotein (NR-53788, BEI Resources, Mannassas, VA, USA) for one hour at 4ºC under constant agitation, before the addition of 1 IU horseradish peroxidase (SERVA GmbH, Heidelberg, Germany) and further incubation for 1h. Unbound sites were blocked with 250 µg BSA for 30 mins before centrifugation at 8,800g for a further 30 mins. The pellet was washed twice and re-spun, prior to final resuspension to OD 50520 nm in 20 µL. The suspension was applied on a conjugate pad, air-dried, and installed in the modified LFD (Innova Medical Group Inc., Pasadena, CA, USA) without the supplied conjugate pad. Serial dilutions of heat-inactivated SARS nCOV-2 in culture media were applied in 100 µL buffer. After 30 minutes, the samples were imaged, followed by the addition of 150 µL DMB Buffer (comprised of 100 µg/mL 3,3'-Diaminobenzidine tetrahydrochloride; Sigma-Aldrich, Gillingham, UK), supplemented with 0.02% v/v hydrogen peroxide and was monitored for color development before final imaging.

**Supplementary Figures**

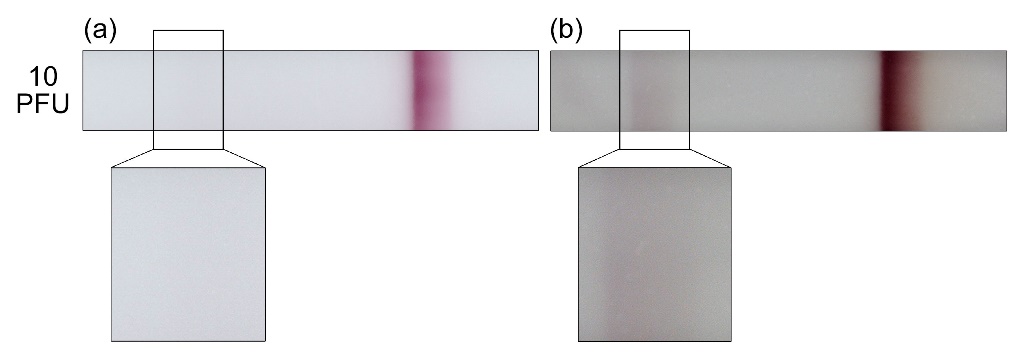

**Supplementary Figure 1.** Use of gold nanoparticles with pre-absorbed mAb anti-SARS nCOV-2 Spike antibodies and Horseradish Peroxidase (HRP). Lateral flow device targeting protein incubated with 10 PFU heat-inactivated viral particles **(a)** and after further development and signal amplification with HRP substrate **(b)**, highlighting the feasibility of amplifying LFD signals. Optical images acquired under identical conditions and at x 20 magnification.

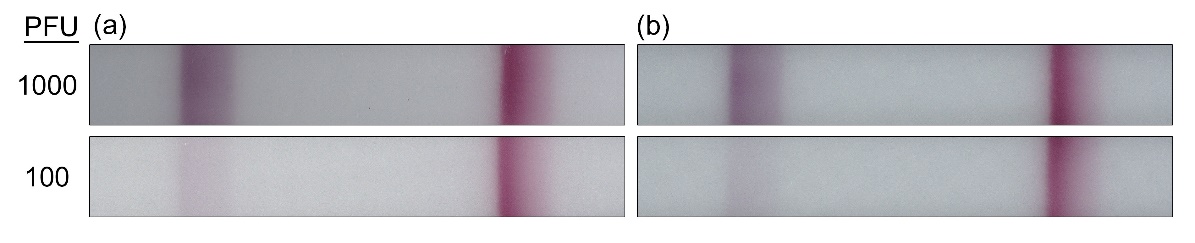

**Supplementary Figure 2.** Comparison between LFDs run using contrived **(a)** or without a biological matrix **(b)** at different PFU concentrations of 1000 (top) and 100 PFU (bottom), demonstrating comparable signal intensity and background color development.
